# Aquaporin-4 as an early cerebrospinal fluid biomarker of Alzheimer’s disease

**DOI:** 10.1101/2024.06.11.24308689

**Authors:** Nerea Gómez de San José, Steffen Halbgebauer, Petra Steinacker, Sarah Anderl-Straub, Samir Abu Rumeileh, Lorenzo Barba, Patrick Oeckl, Giovanni Bellomo, Lorenzo Gaetani, Andrea Toja, Sára Mravinacová, Sofia Bergström, Anna Månberg, Alberto Grassini, Innocenzo Rainero, Peter Nilsson, Lucilla Parnetti, Markus Otto

## Abstract

**Importance:** Aquaporin-4 (AQP4) plays a critical role in the glymphatic system, responsible for clearing brain solutes like Aβ peptides. Exploring AQP4 as an Alzheimer’s disease (AD) biomarker might aid in the understanding of AD neuropathology and monitor the effects of novel drug candidates on the glymphatic system.

**Objective:** To determine the potential of CSF AQP4 as an early stage AD biomarker using a newly established immunoassay.

**Design:** A discovery cohort (n = 157) (2010-2022), composed by AD patients, other neurodegenerative conditions and controls (CON), was used to assess the diagnostic performance of CSF AQP4. Subsequently, AQP4 concentration across the clinical AD spectrum was analyzed in two independent validation cohorts (n = 176) (2016-2023). Stratified randomization based on diagnosis and blinded analyses were performed.

**Setting:** Multicenter study: Ulm University Hospital (discovery), University of Perugia (validation cohort I), University Hospital of Torino (validation cohort II).

**Participants:** Discovery cohort: 38 CON, 40 AD, 21 primary progressive aphasia, 20 behavioural variant frontotemporal dementia, 17 amyotrophic lateral sclerosis (ALS), and 21 Lewy body disease (LBD). Validation cohorts: 55 CON, 14 preclinical AD, 51 AD with mild cognitive impairment (AD-MCI), 39 AD dementia (ADD) and 17 mild cognitive impairment with non-AD pathology (non-AD MCI). The discovery cohort was selected through random sampling, while validation cohort I and II followed a consecutive sampling method.

**Exposures:** CSF AQP4

**Main Outcome (s) and Measure (s):** AQP4 CSF biomarker detection

**Results:** A total of 333 participants were included in this study. In the discovery cohort, the median (IQR) age was 69 (61-75) years and 46.5% of the cohort were women. CSF AQP4 concentration was increased in AD patients compared to CON (p < 0.001), ALS (p = 0.015), and LBD (p = 0.012) patients. CSF AQP4 in AD patients were further analyzed in validation cohort I (median (IQR) age, 74 (71-77) years; 62.0% women), and II (median (IQR) age, 71 (65-75) years; 58.5% women). When analyzing the different stages of the AD continuum in validation cohort I, AD-MCI (p = 0.011) and ADD (p = 0.002) patients had significantly higher AQP4 concentrations than CON. Similar results were obtained in cohort II, where AQP4 levels were higher in AD-MCI (p < 0.001) and ADD (p = 0.028) patients compared to controls. The AQP4 accuracy (area under the receiver operating characteristic curve [AUC]) to distinguish AD patients from CON was 0.81 (95% CI: 0.71 to 0.90, p <0.001) in the discovery cohort, 0.70 (95% CI: 0.60 to 0.81, p<0.001) in validation cohort I and 0.82 (95% CI 0.71 to 0.94, p <0.001) in II. Moreover, patients with AD-MCI could be distinguished from non-AD MCI with an AUC of 0.79 (95% CI: 0.65 to 0.93, p = 0.002).

**Conclusions and Relevance:** Three independent cohorts consistently showed elevated AQP4 levels in AD (including AD-MCI and ADD) compared to CON and other neurodegenerative conditions, suggesting specificity to AD pathology. These findings contribute to understanding AD neuropathology and propose AQP4 as a potential early biomarker of AD. Further investigations are needed to proof AQP4 as a fluid blood brain barrier damage marker.

**KEY POINTS:** *Question:* Is cerebrospinal fluid (CSF) aquaporin-4 (AQP4) an early Alzheimer’s disease (AD) biomarker?

*Findings:* In this cross-sectional study of 333 participants from 3 different cohorts, the CSF concentration of AQP4 was significantly increased in patients with AD, both with mild cognitive impairment (MCI) and dementia, when compared to controls (CON) and other neurodegenerative conditions.

*Meaning:* CSF AQP4 is altered in early stages of AD and might be a fluid biomarker for blood brain barrier damage.

## INTRODUCTION

The misdiagnosis of neurodegenerative diseases poses a significant challenge in the field, with up to 25-30% of patients with Alzheimer’s disease (AD) receiving an incorrect diagnosis ^1^. AD stands as the most prevalent neurodegenerative disorder, accounting for approximately 60-70% of all dementia cases ^1^. Patients diagnosed with AD typically present impaired episodic memory that progresses into other cognitive symptoms, including language difficulties and challenges with executive and visuospatial functions, ultimately leading to the development of dementia ^1^. AD is characterized by two major neuropathological features: extracellular plaques of amyloid-β (Aβ) peptides and intracellular neurofibrillary tangles of hyperphosphorylated tau protein ^2^.

The diagnosis of AD relies on the clinical evaluation of the patient, often complemented by the analysis of three cerebrospinal fluid (CSF) biomarkers. These core biomarkers include the amyloid-β ratio (Aβ42/40), phosphorylated-Tau181 (p-tau) and total-Tau (t-tau) reflecting cerebral amyloidosis, tauopathy and neurodegeneration, respectively ^3^. However, it is clear nowadays that alterations other than Aβ and tau deposition, e.g., synaptic dysfunction blood-brain-barrier (BBB) impairment and impaired protein clearance, may take place in early disease stages, before consistent neurodegeneration occurs. Therefore, additional CSF biomarkers are needed.

Aquaporin-4 (AQP4) is a water channel highly expressed in the central nervous system (CNS) ^4,5^, prominently enriched in the perivascular endfeet of astrocytes wrapping around blood vessels in the periarteriolar and perivenular space ^6,7^. It plays a crucial role in the glymphatic system ^8^, enabling the exchange between CSF and interstitial fluid and supporting the clearance of brain solutes ^9–12^. The impact of AQP4-mediated glymphatic flow on the clearance of solutes may hold clinical relevance, particularly in neurodegenerative diseases characterized by the accumulation of neurotoxic deposits, such as AD ^10,13^. In this regard, *Aqp4* knockout mice models have demonstrated that Aβ and tau are removed from the brain along this paravascular network ^10,14–16^. Moreover, alterations in the expression and localization of this protein have been observed in AD patients ^17^. This change could explain the comprised glymphatic system characteristic of the aging and AD-affected brain ^14,18–21^. However, so far no well validated AQP4 immunoassay is available for the analysis of protein concentrations in CSF.

Herein, we investigate alterations in the concentration of AQP4 protein in CSF of neurodegenerative conditions, with a particular focus on AD. Utilizing a newly developed, highly sensitive AQP4 ELISA, we screened AQP4 concentrations in 157 CSF samples from patients with a broad range of neurodegenerative conditions and controls (CON). Two additional independent cohorts, consisting of 176 CSF samples, were analyzed to further validate the AQP4 results obtained in the AD patient group.

## METHODS

### 1. Participants and clinical characterization

In this project, a discovery cohort of 157 CSF samples gathered at Ulm University Hospital from 2010 to 2022 was analyzed. The cohort comprised 40 AD, 21 primary progressive aphasia (PPA), 20 behavioral variant frontotemporal dementia (bvFTD), 17 amyotrophic lateral sclerosis (ALS), 21 Lewy body disease (LBD) and 38 CON (**Table 1**). To reinforce the robustness of the results, a validation step using two independent cohorts from University of Perugia (validation cohort I) and University of Turin (validation cohort II) was conducted. Those cohorts comprised a total of 176 additional CSF samples: 14 preclinical AD (preAD), 51 mild cognitive impairment due to AD (AD-MCI), 39 AD-dementia (ADD) patients, 17 non-AD MCI and 55 CON (**Table 2**). Diagnosis of the CON participants is described in **Supplementary Table 1**. The study obtained approval from the respective regional ethics committees. All patients provided written informed consent. For further details regarding study design and recruitment procedures, see the eMethods in the Supplement.

**Table 1.** Demographic and clinical characterization of discovery cohort from Ulm University Hospital.

|  |  | CON | AD | PPA<sup>c</sup> | bvFTD | ALS | LBD | Total |
| --- | --- | --- | --- | --- | --- | --- | --- | --- |
| Demographic features | Sample size | 38 | 40 | 21 | 20 | 17 | 21 | 157 |
|  | Sex (F/M) | 21/17 | 21/19 | 8/13 | 6/14 | 9/8 | 8/13 | 73/84 |
|  | Age (years) | 69 (63-74) | 74 (67-79) | 68 (60-74) | 62 (60-67) | 62 (48-72) | 71 (67-75) | 69 (61-75) |
| Cognitive scores | MMSE | NA | 23 (18-26) | 23 (18-25) | 27 (23-28) | NA | NA | 23 (19-27) |
|  | CDR-SOB | NA | 3.5 (2.0-5.5) | 2.0 (1.0-3.5) | 3.0 (2.0-4.0) | NA | NA | 2.8 (2.0-4.5) |
| AD core biomarkers | Aβ42 (pg/ml) | 1266 (873-1493)<sup>a</sup> | 373 (287-527)<sup>b</sup> | 635 (518-1053) | 926 (763-1148) | NA | NA | 707 (433-1088) |
| | Aβ40 (pg/ml) | 13031 (8574-14363)<sup>a</sup> | 10237 (7920-12860)<sup>b</sup> | 10775 (8749-12128) | 10104 (8210-13899) | NA | NA | 10806 (8546-13286) |
| | Aβ42/40 | 0.10 (0.10-0.10)<sup>a</sup> | 0.04 (0.03-0.04)<sup>b</sup> | 0.07 (0.05-0.10) | 0.10 (0.09-0.11) | NA | NA | 0.08 (0.04-0.10) |
|  | t-tau (pg/ml) | 275 (223-346)<sup>a</sup> | 692 (500-1220)<sup>b</sup> | 455 (328-639) | 306 (225-368) | NA | NA | 407 (278-675) |
|  | p-tau (pg/ml) | 37 (30-41)<sup>a</sup> | 103 (76-186)<sup>b</sup> | 62 (36-101) | 38 (27-49) | NA | NA | 51 (35-100) |
|  | A+T+ (%) | 0 | 97.5 | 42.8 | 10 | NA | NA | 41.2 |

Age, MMSE, CDR-SOB, Aβ42, Aβ40, Aβ42/40, t-tau, and p-tau are displayed as median and IQR in brackets. Only AD core biomarkers measured with Lumipulse<sup>®</sup> are displayed in the table. <sup>a, b</sup> 27 CON and 20 AD were analyzed with the ELISA INNOTEST<sup>®</sup> and their values are therefore not included in the table. <sup>c</sup> Within the PPA group, 7 patients presented logopenic variant, 7 semantic variant and 7 non-fluent variant PPA. AD, Alzheimer's disease; ALS, amyotrophic lateral sclerosis; bvFTD, behavioral variant frontotemporal dementia; CDR-SOB, clinical dementia rating-sum of boxes; CON, control; F, female; IQR, interquartile range; LBD, Lewy body disease; M, male; MMSE, mini-mental state examination; NA, not available; PPA, primary progressive aphasia.

**Table 2.** Demographic and clinical characterization of validation cohorts (cohort I and cohort II)

| Cohort I: University of Perugia |  |  |  |  |  |  |
| --- | --- | --- | --- | --- | --- | --- |
|  | Diagnostic group | CON | preAD | AD-MCI | ADD | Total |
| Demographic features | Sample size | 37 | 14 | 28 | 29 | 108 |
|  | Sex (F/M) | 19/18 | 11/3 | 17/11 | 20/9 | 67/41 |
|  | Age (years) | 75 (72-78) | 76 (69-76) | 73 (69-75) | 76 (70-79) | 74 (71-77) |
| Cognitive scores | MMSE | 27 (23-28) | 27.5 (27-28) | 24 (21-26) | 16 (11-21) | 24 (19-27) |
|  | CDR | 0.5 (0-0.5) | 0 (0-0.5) | 0.5 (0.5-0.5) | 1 (1-2) | 0.5 (0.5-1) |
| AD core biomarkers | Aβ42 (pg/ml) | 1011 (753-1355) | 647 (565-832) | 633 (489-786) | 537 (409-822) | 724 (538-1009) |
| | Aβ40 (pg/ml) | 10815 (8329-12923) | 14399 (9262-15958) | 14453 (10198-16904) | 11953 (8181-15408) | 11850 (8946-15679) |
| | Aβ42/40 | 0.097 (0.092-0.106) | 0.056 (0.049-0.063) | 0.045 (0.040-0.060) | 0.049 (0.046-0.058) | 0.06 (0.047-0.093) |
|  | t-tau (pg/ml) | 260 (205-313) | 432 (341-639) | 705 (543-898) | 731 (576-1006) | 536 (294-765) |
|  | p-tau (pg/ml) | 34 (28-43) | 69 (59-100) | 90 (71-119) | 91 (70-128) | 69 (40-100) |
|  | A+T+ (%) | 0 | 100 | 100 | 100 | 65.7 |
| Cohort II: University of Turin |  |  |  |  |  |  |
|  | Diagnostic group | CON | AD-MCI | ADD | non-AD MCI | Total |
| Demographic features | Sample size | 18 | 23 | 10 | 17 | 68 |
|  | Sex (F/M) | 8/10 | 15/8 | 8/2 | 9/8 | 40/28 |
|  | Age (years) | 68 (62-75) | 73 (69-74) | 72 (62-77) | 70 (63-74) | 71 (65-75) |
| Cognitive scores | MMSE | 28 (27-29) | 22 (21-24) | 16 (7-23) | 26 (24-28) | 25 (21-27) |
| AD core biomarkers | Aβ42 (pg/ml) | 987 (654-1384) | 463 (294-523) | 317 (231-394) | 548 (396-972) | 508 (327-743) |
| | Aβ40 (pg/ml) | 9846 (7035-13383) | 10289 (7410-11186) | 7726 (6870-8563) | 6526 (4187-9202) | 8209 (6198-11109) |
| | Aβ42/40 | 0.098 (0.090-0.108) | 0.043 (0.038-0.048) | 0.039 (0.033-0.052) | 0.091 (0.078-0.104) | 0.069 (0.042-0.096) |
|  | t-tau (pg/ml) | 192 (79-274) | 795 (532-1004) | 740 (432-1357) | 319 (174-512) | 414 (202-797) |
|  | p-tau (pg/ml) | 28 (21-40) | 118 (74-184) | 128 (82-190) | 38 (19-103) | 66 (30-138) |
|  | A+T+ (%) | 0 | 100 | 90 | 0 | 47 |

Age, MMSE, CDR, Aβ42, Aβ40, Aβ42/40, t-tau, and p-tau are displayed as median and IQR in brackets. AD, Alzheimer's disease; ADD, Alzheimer's disease dementia; CDR, clinical dementia rating; CON, control; F, female; IQR, interquartile range; M, male; MCI, mild cognitive impairment; MMSE, mini-mental state examination; preAD, preclinical AD.

### 2. Sample collection and biomarker measurements

CSF was obtained through lumbar puncture and centrifuged between 2000-2500 g for 10 minutes at 4°C. The resulting supernatants were aliquoted and promptly stored at −80°C no later than 2 hours after collection. CSF AD core biomarkers (Aβ42, amyloidβ 1-40 peptide (Aβ40), p-tau, and t-tau) were analyzed with Lumipulse® G600-II system (Fujirebio Europe, Ghent, Belgium) or ELISA (INNOTEST®, Fujirebio). Stratified randomization was performed based on the clinical diagnosis, and samples were analyzed in a blinded manner. All CSF used for analytical validation and as QC samples were remnant samples of healthy donors from Ulm University Hospital. Samples were thawed at room temperature (RT), vortexed, centrifuged at 10,000 g for 5 min at 4°C, and kept on ice until measurement.

### 3. AQP4 ELISA

The AQP4 ELISA was developed with one polyclonal (Cell signaling, 59678BF) and one monoclonal (Abcam, ab248213) antibody targeting the C-terminal region of the AQP4 protein, which is located on the intracellular side of the plasma membrane. The calibrator used in the ELISA (62.5-3500 pg/ml) was an AQP4 fusion protein (Proteintech, Ag9561) containing a region of the C-terminal intracellular domain (T_208_-V_323_, UniProt identifier (P55087). The fusion protein contained a GST Tag and was expressed using the bacterial vector PGEX-4T on *E.coli*. The full ELISA procedure can be found in the Supplements.

The ELISA assay was analytically validated for CSF applications following the previously described guidelines ^22^. The following parameters were analyzed: limits of quantification, precision, parallelism, spike-recovery, dilutional linearity, sample stability, and specificity. For further details on the analytical validation, see eMethods in the Supplements.

### 4. Antibody-based suspension bead array assay

A suspension bead array assay was performed following the previously published protocol ^23–25^. In brief, four different antibodies targeting AQP4 were coupled onto color-coded magnetic beads, followed by incubation with diluted CSF samples labelled with biotin. A streptavidin-conjugated fluorophore enabled read-out using a Flexmap 3D instrument (Luminex corporation). For more details see supplemental information.

### 5. Statistical analyses

Data processing, analysis, and visualization were conducted using R (version 4.2.2, R Foundation for Statistical Computing) and GraphPad Prism (version 8.3.0, La Jolla, California, USA). R packages dplyr, tidyverse, ggplot2, ggpubr, ggbeeswarm, emmeans, dunn.test, and pROC were employed. Multiple linear regression models, incorporating age, and sex as covariates, assessed the impact of clinical diagnosis on CSF AQP4 concentrations. A stepwise backward elimination method identified key variables. Assumptions of linearity, homoscedasticity, and normality of residuals were verified for model validity. To address skewed and non-normally distributed biomarker concentration data, a logarithmic transformation was applied before conducting further analysis. Tukey’s test facilitated pairwise comparisons of estimated marginal means. Univariate analysis involved Shapiro-Wilk and Levene’s tests for normality and variance homogeneity. Bonferroni correction was employed for multiple testing. Two-sided hypothesis testing was applied throughout the analyses. Non-parametric Spearman’s correlation were analyzed. Receiver operating characteristics (ROC) curves illustrated sensitivity and specificity at various cut-off points. Significance levels were denoted as *p < 0.05, **p < 0.01, and ***p < 0.001.

## RESULTS

An ELISA assay was used to determine the AQP4 concentrations in CSF samples from individuals with neurodegenerative diseases. A focus on AD aimed to investigate alterations in this protein during the early stages of the disease and determine its diagnostic potential.

### 1. AQP4 ELISA development and performance

The ELISA assay successfully passed all the analytical validation tests for precision, parallelism, spike-recovery, dilutional linearity, and protein stability ^22^ (see eMethods). The assay detected AQP4 in CSF samples with an LLOQ of 127 pg/ml based on calculations with the blocking buffer as blank. Repeatability and intermediate precision were assessed for three QC samples with high, medium, and low levels of AQP4, ranging from 2.6% to 4.5% and 12.9% to 16.3%, respectively (**Supplementary Table 2**). The specified acceptance criteria (75-125%) for parallelism were successfully fulfilled, with a minimum required dilution of 1/4 (**Supplementary Figure 1A**). Moreover, all subsequent dilutions remained within the acceptable range, demonstrating parallelism throughout the experiment. Furthermore, the binding characteristics of the antibodies to the endogenous analyte in the biological matrix closely resembled those observed for the recombinant protein in the blocking buffer (**Supplementary Figure 1B**). Moreover, the criteria were effectively met for spike-recovery (recovery percentage 95-107%) (**Supplementary Table 3**) and dilutional linearity (**Supplementary Figure 1C**). Last, AQP4 was stable for up to five freeze-thaw cycles and when stored at 4°C and RT for up to five days (**Supplementary Figure 1D-F**).

To add additional evidence to the correct target binding, we employed a previously published method ^26,27^ and developed a multiplexed antibody-based suspension bead array assay using four distinct AQP4 antibodies (HPA014784, HPA014944, PAB20767 and 16473-1-AP). The data derived from the bead array and the ELISA described a strong correlation for all antibodies, thereby supporting the specificity of the ELISA method (**Supplementary Figure 2**). Additionally, to further assess the assay’s specificity, CSF samples were spiked with the most abundant proteins in CSF (HSA and IgGs) ^28^. No cross-reactivity between the antibodies used in the assay and HSA or IgG was observed (see eMethods).

### 2. Discovery cohort: AQP4 in neurodegenerative conditions

The initial screening was performed on a cohort of 157 clinical samples, which included 40 AD, 21 PPA, 20 bvFTD, 17 ALS, 21 LBD, and 38 CON (**Table 1**).

A multiple linear regression model was fitted to estimate the relationship between the clinical diagnosis and CSF AQP4 concentration while accounting for age and sex. Among the covariates analyzed, only age (log-transformed β_age_ = 0.013, p < 0.001) (**Supplementary Table 4**) exhibited a significant association with CSF AQP4 concentrations (**Figure 1**) and was therefore included in the model.

**Figure 1.**
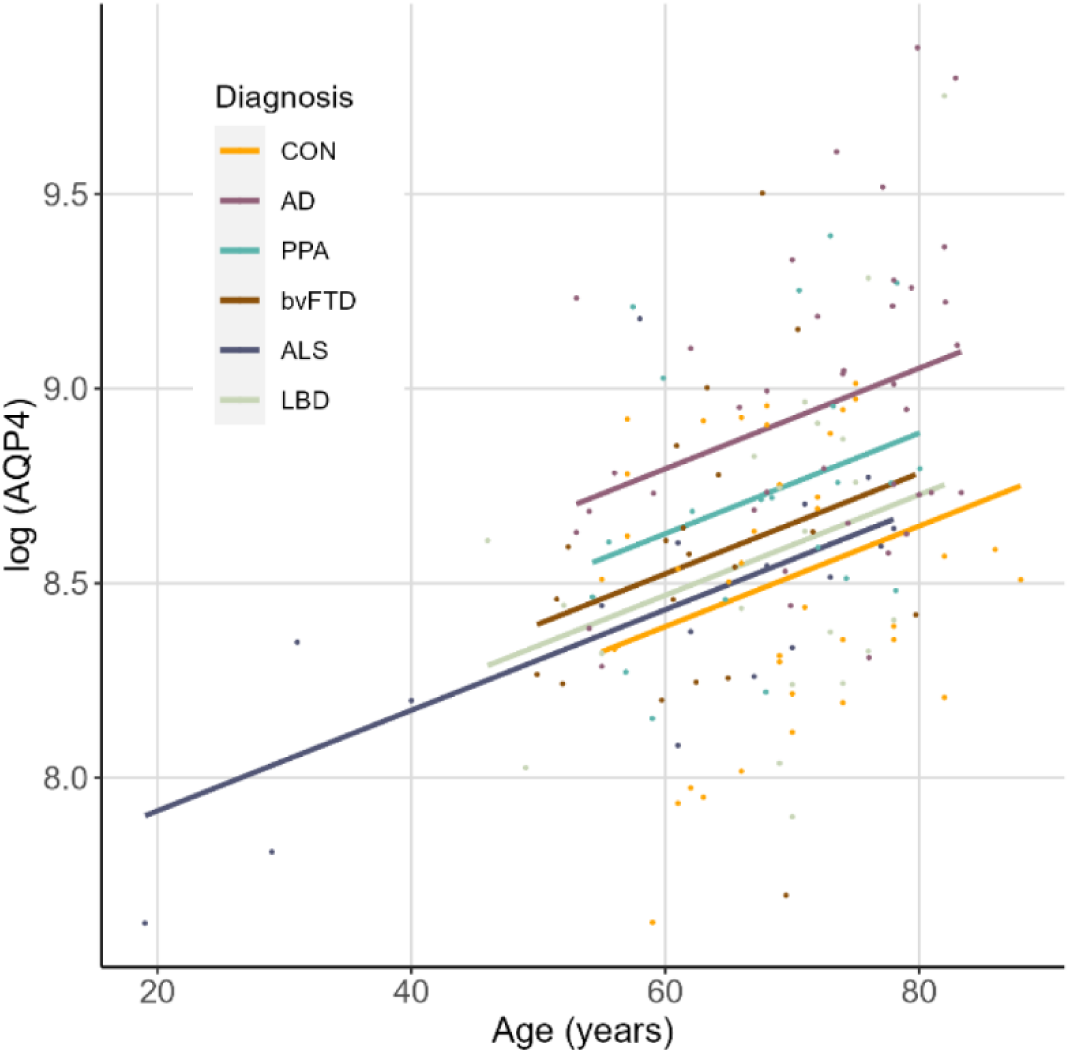
Multivariable regression model. Visualization of the multivariable regression model with diagnosis as independent variable and age as covariate. Y = βCON + βDiagnosis(Diagnosis) + Xage βage + ε. AD, Alzheimer’s disease; ALS; amyotrophic lateral sclerosis; AQP4, Aquaporin-4; bvFTD, behavioral variant frontotemporal dementia; CON, control; LBD, Lewy body disease; PPA, primary progressive aphasia.

The analysis revealed a significant relationship between CSF AQP4 concentration and diagnosis (p < 0.001). Furthermore, the post hoc analysis (Tukey’s Test) indicated higher AQP4 concentration in AD patients than in CON (p < 0.001), ALS (p = 0.015), and LBD (p = 0.012) patients (**Figure 2A**). ROC analysis was conducted to evaluate the diagnostic performance of CSF AQP4 in distinguishing AD patients from CON. The area under the curve (AUC) for the AD vs. CON discrimination was 0.81 (95% CI: 0.71 to 0.90, p <0.001) (**Figure 2B**).

**Figure 2.**
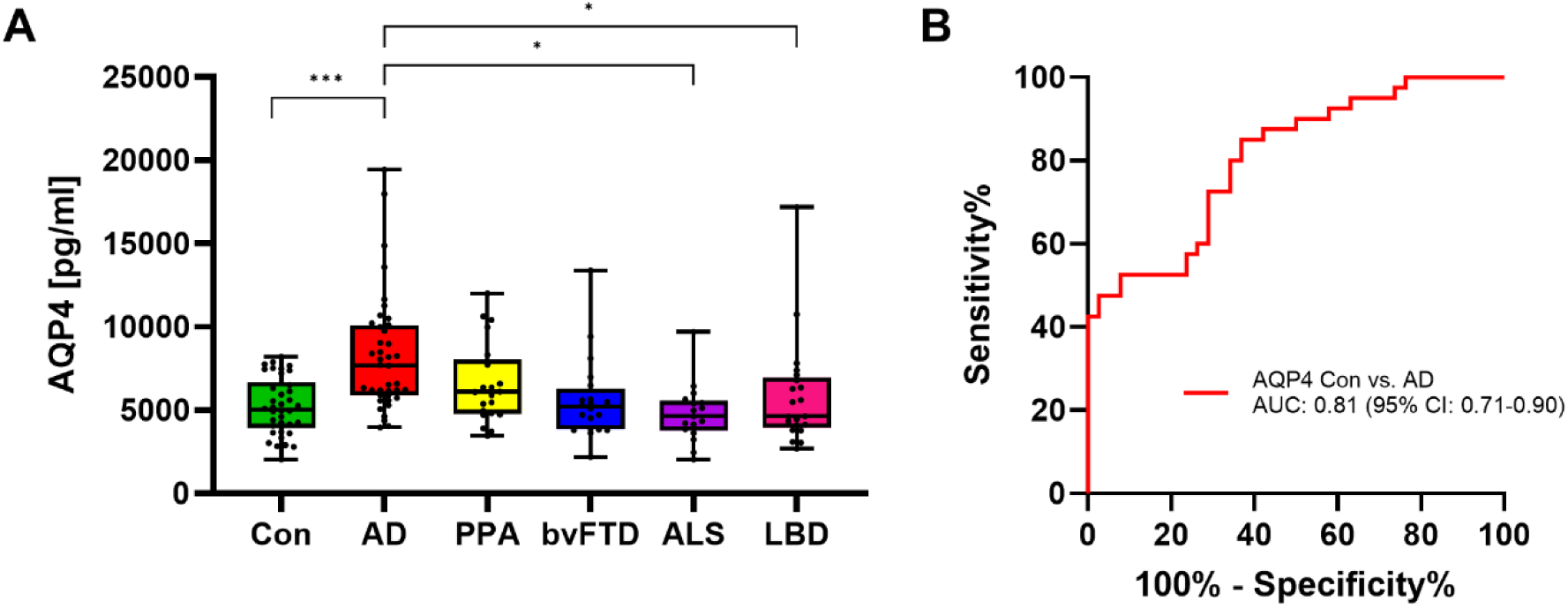
Analysis of CSF AQP4 in the discovery cohort (n = 157) including 38 CON, 40 AD, 21 PPA, 20 bvFTD, 17 ALS and 21 LBD. (**A**) CSF AQP4 concentrations obtained by ELISA in the diagnostic groups. The boxplots represent the median concentration, Q1 and Q3 quartiles, and whiskers from the minimum to the maximum values. A linear regression model was performed, followed by a post hoc multiple comparison test (Tukey’s test) of the estimated marginal means. The levels of statistical significance were set as follows: *p < 0.05, **p < 0.01, and ***p < 0.001. (**B**) ROC curve analysis of CSF AQP4 in AD patients compared to CON. AD, Alzheimer’s disease; ALS; amyotrophic lateral sclerosis; AUC, area under the curve; AQP4, Aquaporin-4; bvFTD, behavioral variant frontotemporal dementia; CON, control; CSF, cerebrospinal fluid; IQR, interquartile range; LBD, Lewy body disease; PPA, primary progressive aphasia; ROC, receiver operating characteristics.

### 3. Validation cohorts I and II: AQP4 in the AD continuum

To validate the findings within AD, the CSF levels of AQP4 were further measured in two independent validation cohorts (**Table 2**). In both cohorts, no age differences were observed between the clinical groups, except for the AD-MCI patients of validation cohort I, which were younger than the CON (p = 0.022). Moreover, the covariates analyzed (age and sex) were not significant in the models.

Validation cohort I included patients with preAD, AD-MCI and ADD in addition to CON subjects. The univariate analyses revealed higher AQP4 protein concentrations in CSF of AD-MCI (p = 0.011) and ADD (p = 0.002) patients than in CON (**Figure 3A**). The ROC analysis revealed the best performance for the comparison CON vs ADD with an AUC of 0.74 (95% CI: 0.61 to 0.86, p 0.001) (**Figure 3B**).

**Figure 3.**
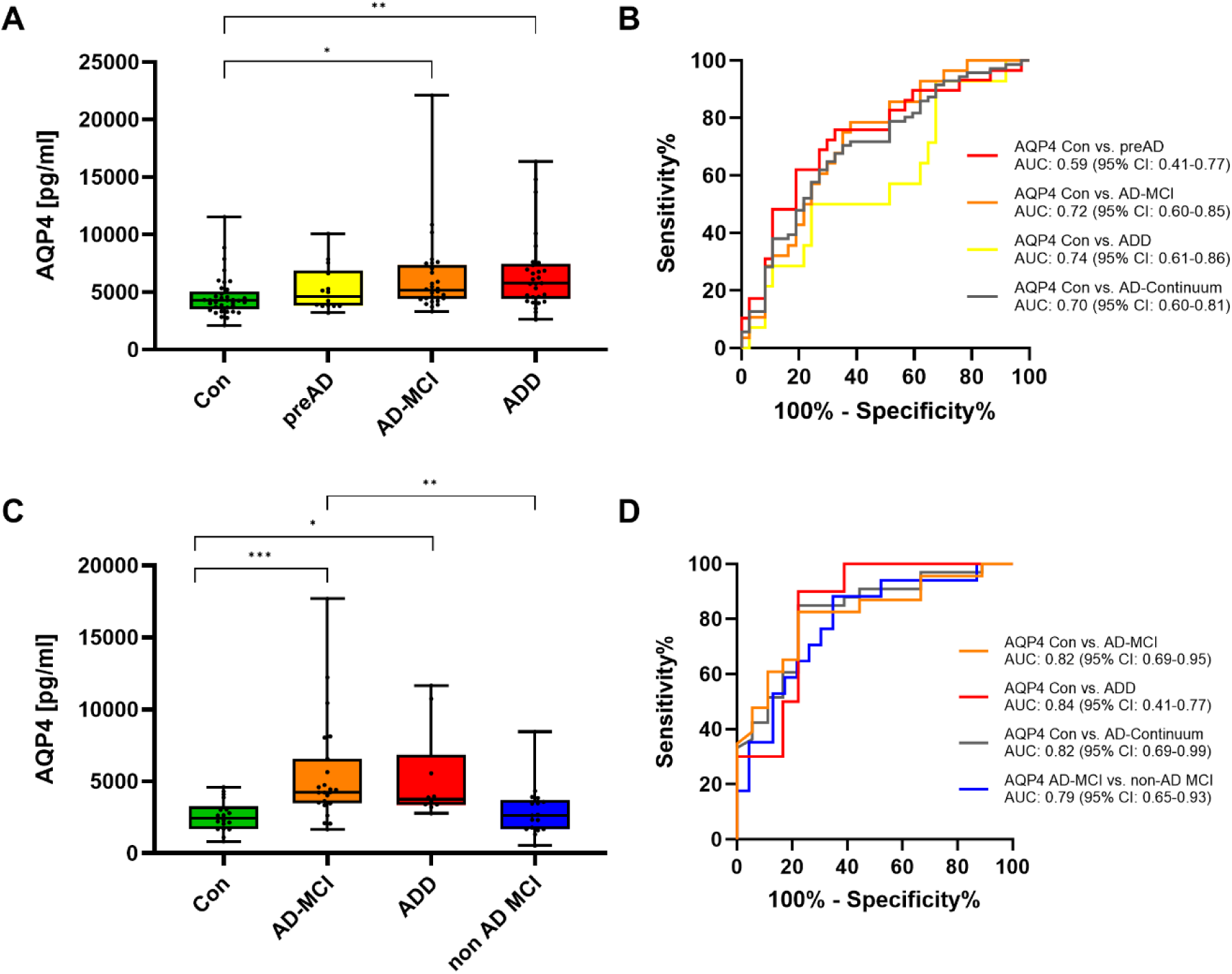
Analysis of CSF AQP4 in validation cohort I and II: cohort I (n = 108) including 37 CON, 14 preAD, 28 AD-MCI and 29 ADD (**A, B**) and cohort II (n = 68) including 18 CON, 23 AD-MCI, 10 ADD and 17 non-AD MCI (**C, D**) (**A**) CSF AQP4 concentrations obtained by ELISA in the diagnostic groups of cohort I (**B**) ROC curve analysis of CSF AQP4 in cohort I (CON vs. AD continuum, preAD, AD-MCI, and ADD). (**C**) CSF AQP4 concentrations obtained by ELISA in the diagnostic groups of cohort II (**D**) ROC curve analysis of CSF AQP4 in cohort I (CON vs. AD continuum, AD-MCI, ADD, non-AD MCI). The boxplots represent the median concentration, Q1 and Q3 quartiles, and whiskers from the minimum to the maximum values. A Kruskal-Wallis followed by Dunn’s Test (with Bonferroni correction for multiple comparisons) was performed. The levels of statistical significance were set as follows: *p < 0.05, **p < 0.01, and ***p < 0.001. AD, Alzheimer’s disease; ADD, Alzheimer’s disease dementia; AQP4, Aquaporin-4; AUC, area under the curve; CI, confidence interval; CON, control; CSF, cerebrospinal fluid; IQR, interquartile range; MCI, mild cognitive impairment; preAD, preclinical AD; ROC, receiver operating characteristics.

Validation cohort II included individuals with AD-MCI, ADD, CON, in addition to a group of non-AD MCI participants. Significantly increased levels of CSF AQP4 were observed in the AD-MCI (p < 0.001) and ADD (p = 0.028) patients compared to CON (**Figure 3C**). In addition, the group of individuals with AD-MCI has significantly higher levels than those with non-AD MCI (0.004) (**Figure 3C**). Furthermore, the ROC analyses revealed the following AUCs: 0.82 (CON vs AD continuum) (95% CI: 0.71 to 0.94, p <0.001), 0.82 (CON vs AD-MCI) (95% CI: 0.69 to 0.95, p <0.001), 0.84 (CON vs ADD) (95% CI: 0.69 to 0.99, p = 0.003) and 0.79 (AD MCI vs non-AD MCI) (95% CI: 0.65 to 0.93, p = 0.002) (**Figure 3D**).

### 4. Correlation with markers of tau pathology, neurodegeneration and cognition

Subsequently, the association between the concentrations of AQP4 and the core biomarkers t-tau and p-tau was investigated among individuals with AD, regardless of their disease stage (preAD, AD-MCI or ADD), across the three cohorts under analysis (discovery, validation cohort I and II) (**Supplementary Figure 4**). Notably, in the discovery cohort, a moderate to strong correlation between AQP4 and both p-tau (r=0.7 (95% CI: 0.35-0.87), p<0.001) and t-tau (r=0.75 (95% CI: 0.44-0.90), p<0.001) was observed in AD patients (**Supplementary Figure 4A**). This correlation persisted across the entire cohort (**Supplementary Figure 4B**). Similar trends, although with comparatively weaker correlations, were delineated in both validation cohort I and Cohort II (**Supplementary Figure 4C-F**).

Furthermore, we assessed the association between CSF AQP4 levels and cognitive decline by correlation with the mini mental state examination (MMSE). We found negative spearman r values in all three cohorts which, however, was only significant in validation cohort II with an r of −0.48 (−0.65 - −0.27), p<0.0001).

## DISCUSSION

Our study aimed to determine the concentration of AQP4 in CSF of AD patients assessing if altered AQP4 levels are a feature of AD also in CSF. The successfully developed and analytically validated ELISA revealed higher concentrations of AQP4 in AD than in control patients, a pattern replicated in two validation cohorts. These findings supported the previously published data using a semi-quantitative antibody-based suspension bead array assay ^27^. To our knowledge, this is the first immunoassay able to perform absolute AQP4 quantification in CSF samples. A recent publication using a commercial AQP4 ELISA reported increased AQP4 levels in the CSF of AD patients ^29^. However, we were unable to identify any signal in the cerebrospinal fluid using this commercial assay.

A stratification by the degree of cognitive impairment of the AD group revealed elevated AQP4 in individuals with MCI, as previously described by Bergström et al. in MCI patients with abnormal tau levels ^27^. Moreover, the changes in MCI seem to be specific to AD as patients with MCI not related to AD pathology had AQP4 concentrations similar to CON. In addition, AQP4 correlated with biomarkers of tau pathology (p-tau) and neurodegeneration (t-tau), as observed for other brain-enriched proteins ^30^.

Our findings in AD patients align with prior studies in human *post-mortem* brain tissue that describe increased AQP4 immunoreactivity as a feature of AD ^17,31,32^. In addition, two independent studies reveal several single nucleotide polymorphisms in the AQP4 gene as modulators of cognitive decline in AD and the relationship between Aβ burden and sleep quality ^33,34^. However, it is worth noting that AQP4 expression might already be increased in the aging brain, as reported by previous research ^17^. Nevertheless, multiple linear regression analyses revealed that increased AQP4 concentration was significantly associated with AD, independent of age, confirming the findings published by Zeppenfeld et al. ^17^.

Alterations in AQP4 polarization within the astrocytic endfeet are observed in frontal and temporal cortex of patients with AD ^17,35^. Elevated levels of AQP4 in CSF may represent a compensatory mechanism, attempting to restore the loss of AQP4 in the astrocytic endfeet through overexpression. This overexpression may aim at restoring the glymphatic flow, affected in neurodegenerative conditions ^14,18,20,21,36^. Additionally, elevated CSF AQP4 levels in AD patients ^37–39^ might be linked to changes in astrocytic reactivity, a characteristic of neurodegenerative diseases ^40,41^.

While existing CSF biomarkers contribute to AD diagnosis (Aβ42/40, t-tau, p-tau) ^3^, AQP4 could assist on the earlier diagnosis of the disease and reflect additional pathological changes still not fully understood. This is supported by the increased AQP4 concentrations in MCI patients. However, no significant differences in AQP4 concentrations were observed in preAD patients, possibly due to the smaller sample size of this group. The potential of AQP4 as an early biomarker may arise from the failure of the glymphatic clearance proceeding Aβ pathology ^17,18^, although conflicting studies suggest that changes in AQP4 expression and localization occur after the formation of the first Aβ plaques ^31,42^. Hence, further studies are needed to determine if AQP4 changes precede protein aggregation and neurodegeneration.

Furthermore, AQP4 emerges as an additional marker to the traditional glial fibrillary acidic protein (GFAP) which shows promise as blood astrocytic injury marker ^43^. The limitations of GFAP as a differential diagnostic marker and its poor protein stability in CSF ^44^, along with overlapping levels between the different diseases and controls ^45^, emphasize the need for additional biomarkers to assess astrocytic status. Additionally, there is no fluid biomarker to address the status of the blood-brain barrier (BBB), and MRI techniques are required to analyze the BBB integrity ^46^. AQP4 could be a potential biomarker for BBB impairment due to the high astrocyte expression surrounding the blood vessels and the required astrocytic polarity for the right BBB permeability ^47^. However, further analysis is needed to test this hypothesis.

Nonetheless, it is essential to recognize and address certain limitations within this study. The cross-sectional study design poses certain constraints, emphasizing the need for longitudinal assessments to evaluate the temporal dynamics of AQP4 as a progression marker. Despite the identification of a significant difference in AQP4 levels between AD and CON, the overlap in protein levels among the diagnostic groups underscores the limitation of AQP4 as a single diagnostic marker. The interpretation of the data is susceptible to potential confounding factors, such as disparate inclusion criteria and recruitment strategies between the cohorts. Moreover, the limited sample size in certain diagnostic groups may also pose limitations. Lastly, the use of different techniques (Lumipulse® and INNOTEST®) for measuring AD core biomarkers in the discovery cohort may introduce inconsistency in data collection.

In conclusion, our study highlights AQP4 as a promising candidate for AD diagnosis and underscores its potential as an early stage biomarker in AD. These findings advocate for further research to ascertain the diagnostic potential and the possible use as an objective readout of therapeutic effects in clinical trials and the use of AQP4 as a fluid biomarker for BBB damage. Furthermore, AQP4 protein levels could help in determining the role of astrocytes and the glymphatic system in AD. Additionally, the development of an AQP4 blood assay could broaden its applicability in the clinical workup.

## Data Availability

All data produced in the present study are available upon reasonable request to the authors

## ACKNOWLEDGEMENTS

[Dum]

## Author contributions

Dr. Otto had full access to all of the data in the study and take responsibility for the integrity of the data and the accuracy of the data analysis

Concept and design: Otto, Halbgebauer

Acquisition, analysis, or interpretation of data: Gómez de San José, Halbgebauer, Otto Drafting of the manuscript: Gómez de San José, Halbgebauer, Otto

Critical revision of the manuscript for important intellectual content: Gómez de San José, Halbgebauer, Steinacker, Anderl-Straub, Abu Rumeileh, Barba, Oeckl, Bellomo, Gaetani, Toja, Mravinacová, Bergström, Månberg, Grassini, Rainero, Nilsson, Parnetti, Otto

Statistical analysis: Gómez de San José, Halbgebauer

Obtained funding: Otto

Administrative, technical, or material support: Rainero, Nilsson, Parnetti, Otto

Supervision: Otto, Halbgebauer

