## Supplements for "Aquaporin-4 as an early cerebrospinal fluid biomarker of Alzheimer’s disease"

### Supplementary Material

#### eMethods

#### Human participants and clinical characterization

*Discovery cohort: Ulm University Hospital*

The discovery cohort was collected at the Neurology department of Ulm University Hospital, Germany, between 2010 and 2022. The cohort included 157 participants from the following diagnostic groups: 40 Alzheimer's disease (AD), 21 primary progressive aphasia (PPA), 20 behavioral variant frontotemporal dementia (bvFTD), 17 amyotrophic lateral sclerosis (ALS), 21 Lewy body disease (LBD) and 38 controls (CON) (**Table 1**). Patients were diagnosed according to international guidelines for disease diagnosis [1–6]. AD patients were checked for AD core biomarkers. The cut-off values for AD core biomarkers measured with Lumipulse® G600-II system (Fujirebio Europe, Ghent, Belgium) were 0.07, 400, and 60 pg/ml for Aβ42/40, t-tau and p-tau, respectively. Moreover, the cut-offs used for ELISA (INNOTEST®, Fujirebio) measurements were 550, 300, and 62 pg/ml for amyloid-β 1-42 peptide (Aβ42), t-tau and p-tau, respectively. The CON participants were analyzed for any sign of neuroinflammation or neurodegeneration by CSF analysis of cell count, total protein, lactate, immunoglobulins quotient, albumin quotient (Qalb), and oligoclonal IgG bands. Furthermore, only individuals who did not exhibit any pathological findings in their cranial magnetic resonance imaging were included in the study (**Supplementary Table 1**). The CON presented no significantly age difference with the rest of diagnostic groups, with an age ranging from 55 to 88 years.

*Validation Cohort I: University of Perugia*

Validation Cohort I consisted of CSF samples collected at the Neurology department of the University of Perugia, Italy, between 2016 to 2023. This cohort included 108 participants: 14 preclinical AD (preAD), 28 AD with mild cognitive impairment (MCI), 29 AD with dementia (ADD) patients and 37 CON (**Table 2**). AD patients underwent a comprehensive neurophysiological evaluation, lumbar puncture, and brain imaging as part of the diagnostic pipeline at baseline. The final AD diagnosis was based on the CSF AD core biomarker profile. The cut-offs used were 0.069, 404 pg/ml, and 56.5 for Aβ42/40, t-tau, and p-tau, respectively. Based on the neurophysiological evaluation and the clinical dementia rating (CDR) scores, the AD patients were classified into three groups: (1) pre-AD patients with a subjective cognitive complaint who did not meet the criteria for MCI; (2) AD-MCI, patients with a clinical dementia rating (CDR) score of 0.5; and (3) ADD patients with a CDR score of ≥ 1.0. All CON participants were subjected to a lumbar puncture to rule out a neurodegenerative condition and presented normal values of AD core biomarkers. Fourteen CON participants reported a subjective cognitive decline but did not meet the MCI criteria. Eleven CON participants were affected by non-degenerative neurological conditions (**Supplementary Table 1**). None of the CON participants showed cognitive impairment after at least 2 years of follow-up. No age difference was observed between the different diagnostic groups, except AD-MCI which were significantly younger ($p = 0.022$) than CON.

*Cohort II: University of Turin*

Cohort II was collected at the Aging Brain and Memory Clinic of the Department of Neuroscience of the University Hospital "Città della Salute e della Scienza" of Turin, Italy. All the 87 patients were recruited consecutively among those who underwent lumbar puncture between the 1st of April 2021 and the 1st of April 2023. The cohort comprises 33 patients with AD, including 23 at MCI stage and 10 at the dementia stage. Additionally, it comprises 17 MCI patients with suspected non-Alzheimer pathology (non-AD MCI) (**Table 3**). The MCI patients with suspected non-Alzheimer pathology include 1 patient fulfilling the diagnostic criteria for normal-pressure hydrocephalus. The control group consisted of 18 cognitively unimpaired individuals (**Supplementary Table 1**). All those 18 individuals showed values of Aβ42/40 ratio within normal limits. The cut-offs used were 0.063, 450 pg/ml, and 60 for Aβ42/40, t-tau, and p-tau, respectively. The diagnoses of AD and suspected non-Alzheimer pathology were made according to Jack et al [7] and the other most recent internationally approved criteria for neurocognitive disorders [3,4,8–12]. Patients with MCI were distinguished from demented patients according to Alberts et al [13]. MCI patients showed a significant cognitive impairment at the neuropsychological examination, but no significant impairment at Activities of Daily living (ADL) [14] and Instrumental Activities of Daily Living (IADL) [15] questionnaires. No statistically significant age differences were observed between the different diagnostic groups.

The studies obtained approval from respective ethics committees (Ulm University Hospital, No. 20/10; University Perugia, 19369/08/AV registry no: 1287/08 date: 9 October 2008 and University of Turin, 0114576) and adhered to the most recent iteration of the Declaration of Helsinki. Prior to their participation, all the participants or their caregivers received a thorough explanation of the study's procedures and gave written consent. The sample size was based on availability.

**Antibody-based suspension bead array assay**

Two polyclonal were selected from the Human Protein Atlas project (www.proteinatlas.org) (HPA014784 and HPA014944) and two from other providers (Abnova, PAB20767 and Proteintech, 16473-1-AP). The antibodies were conjugated onto uniquely color-coded magnetic carboxylated microspheres (MagPlex, Luminex Corporation) after activation of the carboxylic surface with EDC-NHS. Each bead identity corresponded to one antibody. After quenching the remaining bead activity, beads were pooled together into a suspension bead array.

In parallel, CSF samples were labeled with biotin, as previously described [23,24]. Samples were further diluted 1/8 and incubated overnight with the antibody-coupled beads. Antigen targets were cross-linked to the antibody-coated beads using paraformaldehyde. A streptavidin-conjugated fluorophore (Streptavidin R-phycoerythrin conjugate, Invitrogen, diluted to 0.2 µg/ml in PBS 0.05% Tween) was added for the detection of the captured proteins. The Flex-map 3D instrument (Luminex Corporation) was employed for the readout, with median fluorescence intensity (MFI) per bead ID per sample serving as the measurement parameter.

**AQP4 ELISA protocol**

Wells of a 96-well plate (F96 Nunc Immunoplate MaxiSorp, Thermo Fischer Scientific) were coated with 100 µl of 3.3 µg/ml capture antibody (Cell signaling, 59678BF) in coating buffer (100 mM bicarbonate-carbonate buffer pH 9.6) to be adsorbed overnight at 4ºC. The next day, the antibody solution was discarded, and 300 µl of blocking buffer (1% goat serum in PBS 0.05% Tween) were added for 2h at RT. After discarding the blocking buffer, 100 µl of the sample were added and incubated for 1.5 h at 20ºC. The sample solution was discarded, wells were washed 3 times with 300 µl of washing buffer (PBS with 0.05% Tween), and 100µl of 0.66 µg/ml biotinylated detector antibody (Abcam, ab248213) in blocking buffer were applied for 1h at 20ºC. One hour later, the last washing step was conducted, and 100 µL 3,3',5,5'-Tetramethylbenzidin (TMB) (ThermoScientific, 34028) were added to each well and incubated for 30 minutes at RT. The reaction was stopped with 100µL 1M HCl per well. The optical density (OD) was measured at 450nm against a reference wavelength of 570nm with an ELISA reader (Epoch2, BioTek). A four-parameter logistic model (without weighting factor) was fitted to the calibration curve.

**ELISA analytical validation**

The ELISA assay was analytically validated for CSF applications following the previously described guidelines [16]. The following parameters were analyzed: limits of quantification, precision, parallelism, spike-recovery, dilutional linearity, sample stability, and specificity. Sixteen blanks, consisting of the blocking buffer used for each respective assay, were included to calculate the lower limit of quantification (LLOQ).

$$LLOQ = mean + 10 * SD$$

Three CSF samples from single control patients or from pooling different remnants from health donors were used for the analytical testing. For repeatability (intra-assay) and intermediate-precision (interassay), three CSF samples were aliquoted and measured in 5 replicates on 3 non-consecutive days. The precision was expressed in terms of the coefficient of variation (CV%) [16], with a tolerance of 15% for repeatability and 20% for intermediate precision. For parallelism, three CSF samples were serially diluted (two-fold) in blocking buffer over the calibrator range. Both neat and diluted samples were measured in duplicate. The optimal minimum required dilution (MRD) was the sample's lowest dilution, allowing a sample recovery between 75% and 125% for the following dilutions [17]. To assess spike-recovery, CSF samples were diluted to the final dilution used in the assay (MRD or higher dilution), spiked with low, medium, and high concentrations of calibrator antigen (all within the linear range of the calibration curve), and measured in duplicate. Spiked recombinant protein consisted of less than 10% of the total sample volume to avoid further dilution of the sample. 20% variation was accepted for spike-recovery.

$$\text{Spike} - \text{recovery}(\%) = \frac{\text{measured concentration spiked sample}}{\text{measured concentration neat sample} + \text{ theoretical concentration spiked}} \text{ x } 100$$

Moreover, for dilutional linearity, samples were spiked at a concentration 90x above the highest calibration point and diluted 2.5 and 2-fold in blocking buffer to a concentration within the working range. Subsequently, measurements were conducted in duplicate, with an acceptable 15% variation. Protein stability was tested in two CSF samples from control patients. The samples were aliquoted and subjected to 1 to 5 freeze-thaw (FT) cycles or stored under different conditions: at 4ºC for 24h, 3 days, and 5 days; or at RT for 2h, 4h, 24h, 3 days, and 5 days. In all cases, protein stability was determined by comparing the protein concentration to a reference sample stored at -80ºC. Last, to exclude any cross-reactivity with human serum albumin (HSA) and IgG, a physiological and higher concentration of HSA (200 µg/ml and 600 µg/ml) (Sigma, A8763-250MG) and IgG (30 µg/ml and 90 µg/ml) (Sigma, I4506-10MG) was added both in blank and CSF. Moreover, to assess further cross-reactivity with other proteins, an alignment between the epitope of the antibodies (if provided) and the human proteome was performed in NCBI-BLAST ® (https://blast.ncbi.nlm.nih.gov/) [18].

### Supplementary Tables

**Supplementary Table 1**. Diagnosis of the control participants included in the discovery and validation cohorts.

| Diagnosis | n | Cohort |
|---|---|---|
| Abducens nerve paresis | 2 | Discovery cohort Ulm |
| Brainstem transient ischemic attack | 1 | |
| Catatonia | 1 | |
| Epilepsy | 2 | |
| Facial palsy | 4 | |
| Gait disorder, subcortical vascular encephalopathy | 1 | |
| Headache | 5 | |
| Headache, subcortical vascular encephalopathy | 1 | |
| Intracranial hypertension, depression | 1 | |
| Mandibular arthrosis | 1 | |
| Migraine | 1 | |
| Myopathy | 1 | |
| Neuralgic shoulder amyotrophy | 1 | |
| Polyneuropathy | 2 | |
| Post-zoster neuralgia | 1 | |
| Ptosis | 1 | |
| Seizures | 1 | |
| Temporal arteritis (Giant cell arteritis) | 3 | |
| Transient ischemic attack | 1 | |
| Psychiatric disorder | 1 | |
| Trochlear palsy | 3 | |
| Vestibular neuritis | 2 | |
| Vitamin B12 deficiency | 1 | |
| Bipolar disorder | 1 | Validation Cohort I Perugia |
| Cranial neuropathy | 1 | |
| Headache | 1 | |
| Late onset epilepsy | 4 | |
| Melanoma metastasis and epilepsy | 1 | |
| Minimal cognitive impairment | 3 | |
| Multifactorial MCI | 2 | |
| Multifocal ischemic stroke | 1 | |
| Non-neurodegenerative amnesic syndrome | 1 | |
| Normal pressure hydrocephalus | 3 | |
| Paraneoplastic anti-HU polyneuropathy | 1 | |
| Psychiatric disorder | 3 | |
| Stable MCI | 1 | |
| Subjective cognitive decline | 6 | |
| Transient global amnesia | 1 | |
| Vascular dementia | 3 | |
| Vascular MCI | 4 | |
| Autoimmune encephalitis | 1 | Validation Cohort II Turin |
| Myelitis | 1 | |
| Neurologically healthy patient in spinal anaesthesia | 4 | |
| Polyneuropathy | 4 | |
| Subjective cognitive decline | 8 | |

**Supplementary Table 2**. Repeatability and intermediate precision of AQP4 ELISA using three CSF samples with high, medium, and low levels of AQP4 (QC1, QC2, and QC3).

$S_r$ = repeatability standard deviation, $S_{RW}$ = intermediate precision standard deviation

| **Reference sample** | **AQP4 concentration (pg/mL)** | **$S_r$ (pg/ml)** | **Repeatability ($CV_r$%)** | **$S_{Rw}$ (pg/ml)** | **Intermediate precision ($CV_{Rw}$%)** |
|---|---|---|---|---|---|

| | | | | | |
|---|---|---|---|---|---|
| **QC1** | 6526.1 | 169.5 | 2.6 | 1063.1 | 16.3 |
| **QC2** | 3245.4 | 128.3 | 4.0 | 417.9 | 12.9 |
| **QC3** | 1739.0 | 77.9 | 4.5 | 239.3 | 13.8 |
| **Mean** | | | **3.7** | | **14.3** |

**Supplementary Table 3**. Spike-recovery of AQP4 ELISA in three CSF samples.

| | **Recovery percentage (%)** | | |
|---|---|---|---|
| **Sample** | **Low spike (300 pg/mL)** | **Medium spike (800 pg/mL)** | **High spike (1500 pg/mL)** |
| **Sample 1** | 111 | 99 | 96 |
| **Sample 2** | 104 | 105 | 95 |
| **Sample 3** | 106 | 97 | 94 |
| **Mean recovery percentage (%)** | **107** | **100** | **95** |

**Supplementary Table 4**. Summary of the multivariable regression model in the AQP4 discovery cohort.

| | **Estimate** | **Std.Error** | **t value** | **P-value** |
|---|---|---|---|---|
| **Intercept** | 7.613750 | 0.204414 | 37.247 | < 2e-16 |
| **Diagnosis AD** | 0.405436 | 0.080954 | 5.008 | 1.52e-06 |
| **Diagnosis PPA** | 0.238612 | 0.096669 | 2.468 | 0.0147 |
| **Diagnosis bvFTD** | 0.135728 | 0.099853 | 1.359 | 0.1761 |
| **Diagnosis ALS** | 0.043447 | 0.107757 | 0.403 | 0.6874 |
| **Diagnosis LBD** | 0.080369 | 0.096636 | 0.832 | 0.4069 |
| **Age (years)** | 0.012913 | 0.002847 | 4.536 | 1.16e-05 |

Data was log-transformed to reduce skewness and bring closer to a normal distribution. Residual standard error: 0.3553 on 150 degrees of freedom. Multiple R-squared: 0.3054, Adjusted R-squared: 0.2776. F-statistic: 10.99 on 6 and 150 DF, p-value: 3.91e-10

### Supplementary Figures

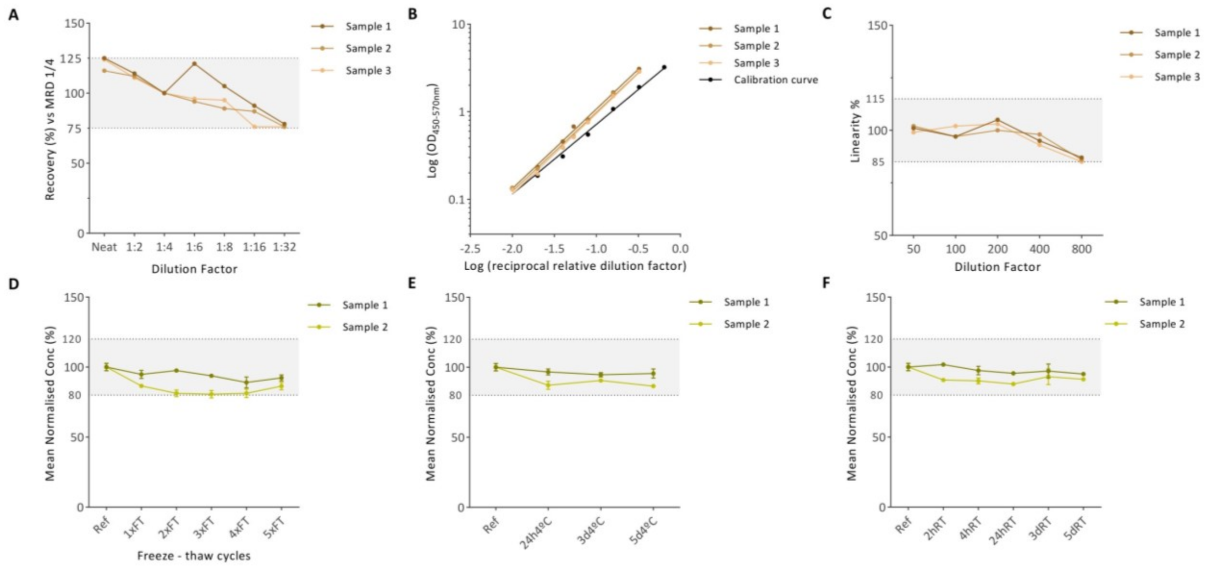


**Supplementary Figure 1.** Analytical validation of the AQP4 ELISA. (**A**) Recovery (%) for each dilution of three CSF samples using 1/4 as the MRD. Recovery (%) was calculated as the ratio between the measured analyte concentration * DF and the measured MRD concentration * DF * 100. (**B**) Parallelism of three CSF samples serially diluted within the quantifiable range of the calibration curve. (**C**) Dilutional linearity of three CSF samples spiked with a high concentration of AQP4 and diluted two-fold into the calibration curve range. Mean percent linearity was calculated as (observed concentration at dilution X) * (DFX) / (observed concentration at dilution X-1) * (DFX-1) * 100. (**D**) Protein stability of AQP4 after multiple FT cycles. The samples were thawed and kept at RT for 4 hours before being frozen at -80°C until analysis. (**E**) Protein stability of AQP4 after keeping the samples at 4°C for 24 hours, 3 days, and 5 days. (**F**) Protein stability of AQP4 after keeping the samples at RT for 2 hours, 4 hours, 24 hours, 3 days, and 5 days until analysis. The horizontal dashed lines represent the accepted variation range. AQP4, Aquaporin 4; CSF, cerebrospinal fluid; DF, dilution factor; FT, freeze-thaw; MRD, minimum required dilution; OD, optical density; Ref, reference; RT, room temperature.

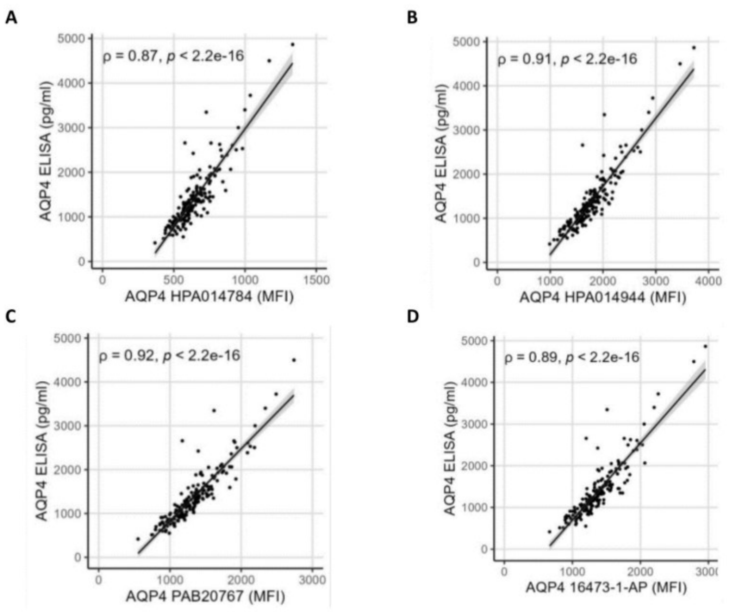


**Supplementary Figure 2.** Correlation between CSF AQP4 levels obtained by ELISA and antibody-based suspension bead array with different AQP4 antibodies: (**A**) HPA014784, (**B**) HPA014944, (**C**) PAB20767 and (**D**) 16473-1-AP in a cohort of 180 participants. Correlation analyses were computed using the Spearman rank correlation coefficient (ρ). The grey area represents the confidence interval. AQP4, Aquaporin-4; CSF, cerebrospinal fluid; MFI, median fluorescent intensity.

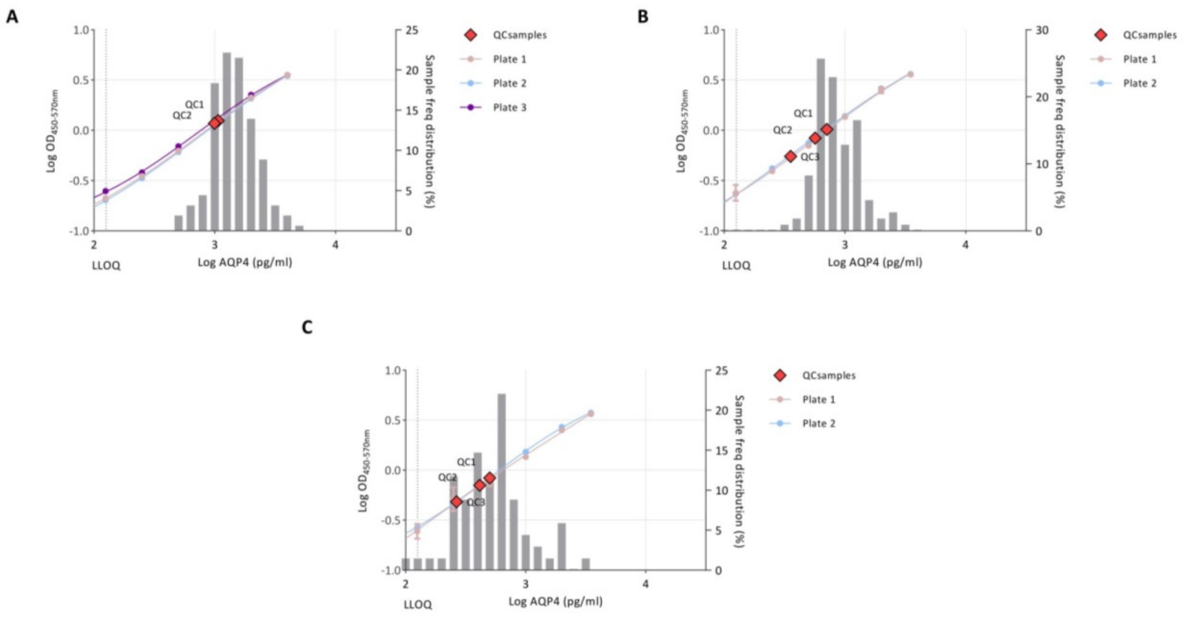


**Supplementary Figure 3.** Frequency distribution of CSF AQP4 levels in the discovery cohort (n = 157) (**A**), cohort I (n = 108) (**B**), and cohort II (n = 68) (**C**) with the calibration curves. Three samples of the discovery cohort and one sample from the cohort I exceeded the calibration curve limit, and their concentrations were extrapolated. One sample from the cohort II was bellow LLOQ. The CSF concentrations of QC samples used in the cohorts are highlighted in red. The vertical dashed line represents the LLOQ. AQP4, aquaporin-4; CSF, cerebrospinal fluid; LLOQ, lower limit of quantification; OD, optical density; QC, quality control.

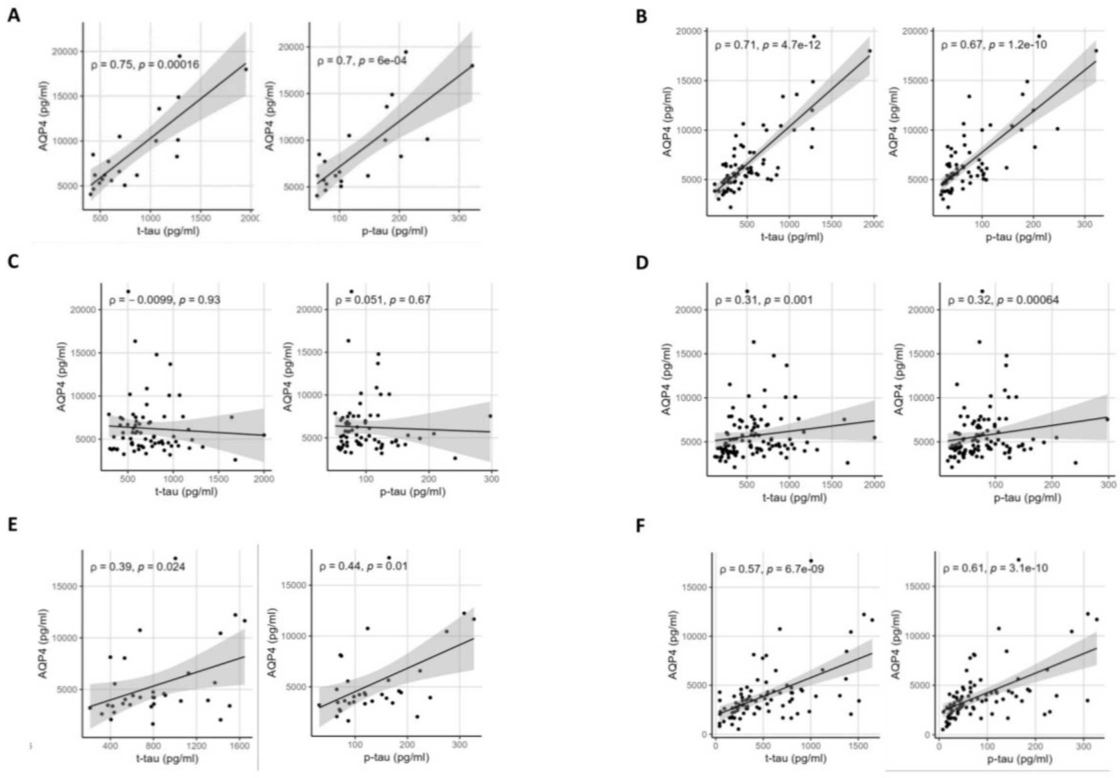


**Supplementary Figure 4**. Correlation of CSF AQP4 with t-tau, p-tau in AD patients (**A**, **C**, **E**) and all diagnostic groups (**B**, **D**, **F**) in the discovery cohort (**A**, **B**), Cohort I (**C**, **D**) and Cohort II (**E**, **F**). Only samples measured with Lumipulse ® were included. Correlation analyses were computed using the Spearman rank correlation coefficient (ρ). The grey area represents the confidence interval. AQP4, aquaporin-4; CSF, cerebrospinal fluid.

**eResults**

**Analytical validation of AQP4 ELISA**

To further analyze off-target antibody binding, a pair-wise sequence alignment was performed between the immunogen sequence of the capture antibody (ab248213) and the human proteome using the NCBI-BLAST ® algorithm [29]. No protein with highly aligned protein sequences was found. The exact epitope of the detector antibody (59678BF) is unknown and the pair-wise sequence alignment for this antibody could not be performed.

**Performance of AQP4 ELISA in Cohort Measurements**

Each cohort was processed on a single day, with samples analysed on parallel ELISA plates. In addition to the clinical samples, three QC samples were measured in triplicate on each plate with a CV% ranging from 2.7% to 18.0%. All AQP4 CSF concentrations were above the LLOQ and fell within the calibrator range (62.5-3500 pg/ml), with the exception of five samples. One sample (cohort II) was below the LLOQ (88 pg/ml), while four (3x discovery cohort and 1x validation cohort I) exceeded the upper limit of the calibration curve and their concentrations were extrapolated (**Supplementary Figure 3**). Furthermore, all calibrator points demonstrated acceptable precision below 20%, (62.5 pg/ml), with a back-calculated concentration ranged from 96% to 115%, except for the lowest calibrator which is below LLOQ.
